# Development and validation of a personalised antipsychotic selection tool for first-line treatment in severe mental illness

**DOI:** 10.1101/2025.03.24.25324323

**Authors:** S Hardoon, DPJ Osborn, M Boman, F Ben Abdesslem, A Richards-Belle, N Launders, K Man, ICK Wong, C Dalman, G Ambler, I Petersen, JF Hayes

**Affiliations:** Division of Psychiatry, UCL, London, UK; North London NHS Foundation Trust, London, UK; Division of Clinical Epidemiology, Karolinska Institute, Stockholm, Sweden; Department of Clinical Neuroscience, Karolinska Institute, Stockholm, Sweden; Research Institutes of Sweden, Stockholm, Sweden; UCL School of Pharmacy, UCL, London, UK; Department of Pharmacology and Pharmacy, University of Hong Kong; Department of Global Public Health, Karolinska Institute, Stockholm, Sweden; Department of Statistical Science, UCL, London, UK; Institute of Epidemiology and Healthcare, UCL, London, UK; Department of Clinical Epidemiology, Aarhus University, Denmark

## Abstract

**Background:** Guidance is lacking on choice of first-line antipsychotic for individuals with incident severe mental illness (SMI). Patients may try several before an effective, well-tolerated drug is identified, delaying symptom improvement. We aimed to develop a personalised selection tool to identify the optimum first-line antipsychotic, based on individual sociodemographic and clinical characteristics.

**Methods:** Risk prediction development and validation study using electronic health records (EHRs) from primary care in England (Clinical Practice Research Datalink) linked to Hospital Episode Statistics, including 11,811 individuals with incident SMI prescribed aripiprazole, olanzapine, quetiapine or risperidone as first-line treatment between 2007-2016. The outcome was time to psychiatric hospitalisation or change to different antipsychotic within 3 years of commencing treatment. Prediction algorithms were developed using Cox proportional hazards models in a 70% training sample and validated in a 30% hold-out sample. This baseline model was compared with machine learning survival models of increasing complexity. Potential predictors included demographics, diagnoses, concomitant medications and laboratory findings.

**Outcomes:** Among 8,225 individuals in the development cohort, 4,456 (54.2%) experienced the outcome. In model validations, 1,022 (53.3%) of 1,916 in the validation cohort did not receive the optimal antipsychotic identified by the personalised selection tool. The predicted 3-year outcome risk if all individuals received the medication assigned by the tool was 6.3% lower (95% CI 4.0%-8.5%) than the observed 3-year risk in the validation cohort, and 10.2% lower (95%CI 7.9%-12.5%) than if individuals were randomly assigned an antipsychotic (corresponding numbers need to treat of 16 and 10). Machine learning approaches did not meaningfully improve model performance.

**Interpretation:** A personalised tool based on EHR data could improve treatment success rates by optimising first-line antipsychotic selection. Machine learning did not outperform traditional prediction methods. Further research will assess the impact on adverse events and in other populations.

**Funding:** UK Research and Innovation grant MR/V023373/1.

**Research in context:** *Evidence before this study:* We searched PubMed for articles published from database inception to December 13, 2024, with no language restrictions. We searched titles and abstracts using the terms ((prediction) AND ((treatment response) OR (treatment rule) OR (treatment outcome)) AND ((psychosis) OR (severe mental illness) OR (schizophrenia) OR (bipolar disorder))). We identified 187 articles for full text screening. A number of studies exist on the prediction of lithium treatment response. A recent systematic review summarised the results of eight studies that used biomarkers, clinical and socio-demographic features to predict treatment response in psychosis, however these commonly compared responders with non-responders, rather than developing treatment selection rules. Two studies did generate treatment selection recommendations. One used a Super Learner in Taiwan National Health insurance data to optimise antipsychotic selection in first episode psychosis, resulting in a 7% improvement in estimated treatment success rate. The second examined antipsychotic selection, choosing between risperidone and aripiprazole, in children using Korean National Health insurance data and found a 1.2-1.5 times increase in antipsychotic continuation using their model compared to their allocated treatment. They found no improvement in performance when comparing machine learning with simple regression models. Neither model has been externally validated. We could not find any models that are in clinical use.

**Added value of this study:** We found that a simple treatment selection prediction model, based on data contained in the electronic health records at the point that an individual with severe mental illness is first prescribed an antipsychotic, could reduce treatment failure rates by 6-10%. In our validation cohort 75% of patients were switched to an alternative antipsychotic medication by the treatment selection tool.

In line with the limited number of previous studies in this area, we did not observe meaningful improvements in predictive properties when machine learning approaches were compared with traditional models.

**Implications of all the available evidence:** Prediction models for optimising treatment selection in psychiatry are becoming increasingly possible with data from electronic health records. Improving treatment selection for people with SMI is low risk, compared to other prediction problems in psychiatry, and could improve long-term outcomes. Models still need full external validation and testing in new cohorts.

## Background

Severe mental illnesses (SMI), including schizophrenia, bipolar disorder and other non-organic psychotic illnesses, have a lifetime prevalence of approximately 1% each, and are in the top twenty diseases in terms of disability. This disability burden has been increasing over the past 30 years.[1] SMI is associated with high levels of premature mortality and morbidity, with a mean age of death 15 years younger than the general population.[2]

SMI is challenging to treat. Thirty different antipsychotics are approved for use in the United Kingdom (UK), but it can take a long time to find the optimal antipsychotic for a patient, delaying potential improvements in symptoms. Early, optimised treatment in incident SMI could reduce symptom burden, non-adherence and morbidity.

Personalised or precision medicine aims to assign a patient the optimal treatment, based on their predicted response; thereby moving from average group outcomes to the prediction of outcomes for individual patients.[3] Algorithms that predict response to different medications based on a patient’s demographic, clinical, genetic and biomarker characteristics may be used to better determine which first-line treatment is most likely to be effective for an individual patient.[4]

Regarding personalising treatment for SMI, there is ongoing work focusing on genome-wide associations[5] and biomarkers (for example, from functional magnetic resonance imaging [6] and electroencephalography[7]). However, these approaches have yet to bear fruit clinically. Despite the rich data available, little attention has been paid to predictive analytics using other data sources such as Electronic Health Records (EHRs), which can provide real-world outcomes and adverse event information for large numbers of patients with SMI. This contrasts with cardiovascular disease (CVD), where algorithms for CVD risk are in use clinically[8]. Models selecting between approved and commonly used treatments are low risk compared to other prediction problems in psychiatry; such as prediction of psychosis or suicide, where false negatives and false positives can have devastating impact [9]. Machine learning procedures may improve on traditional regression methods, because of the ability to discover hidden risk factors and interactions, nonlinear, and higher-order effects, as well as to approximate intricate functions that are poorly represented by individual covariate or interaction terms.[10, 11]

Previous studies have used machine learning methods to predict treatment response in depression,[12, 13] but few personalised medicine studies exist for SMI.[11, 14, 15] The majority of the studies in people with SMI focus on predicting response to particular medications, rather than comparing different medications to obtain an overall personalised treatment selection rule.[16–19] The two previous studies which do, both concern personalised treatment selection for schizophrenia in Asian populations; applicability to other populations and SMI groups is unknown.[20, 21]

The aim of this study was to use a large EHR data set to develop and validate algorithms to predict treatment response to different antipsychotics in incident SMI, in order to identify the optimum first-line antipsychotic treatment. Four antipsychotics, most commonly prescribed as first-line treatments in the UK, were considered: aripiprazole, quetiapine, olanzapine and risperidone.[22] A secondary aim was to compare machine learning methods with traditional regression modelling for development of the algorithms.

## Methods

### Data source

Data came from the Clinical Practice Research Datalink (CPRD) Aurum database, which comprises anonymised EHRs retrieved from participating general practices using EMIS software across the UK [23]. A CPRD data set build from May 2022 was used. The database includes recorded diagnoses, symptoms and other health data (blood tests, health indicators), and all issued medication prescriptions. Medical diagnoses and symptoms information is recorded using a mixture of Read, SNOMED and local EMIS coding systems.

Records from primary care practices in England which had agreed to record linkage were linked by the data providers to secondary care hospital records from Hospital Episode Statistics (HES) data, including records from accident and emergency (A&E) (from 1/4/2007 to 31/03/2020), and admitted patient care (APC) (1/4/1997 to 31/03/2021).

### Study design

In a nationwide retrospective cohort derived from EHRs, we conducted risk prediction algorithm development and validation for predicting response to different antipsychotics as first-line treatment in severe mental illness. We compared traditional risk prediction modelling and machine learning methods.[10, 11]

### Setting

The National Health Service in England.

### Cohort

The cohort comprised of individuals with incident SMI between January 2007 and December 2016, who commenced on one of four antipsychotics as a first-line monotherapy treatment: aripiprazole, quetiapine, olanzapine, and risperidone (identified as the first antipsychotic prescription in prescription records). The 10-year period ensured the newest drug aripiprazole was available in routine practice (following approval in 2004) and the end date enabled three years follow-up (to December 2019) before the 2020 COVID-19 pandemic, and to coincide with the timeframe of HES linkage. Patients aged 10-99 years at time of first prescription were included. Incident SMI cases were defined as the first clinical record of schizophrenia, other psychosis or bipolar disorder. Code lists for these diagnoses were updated from established lists.[24] Further details of how the cohort was defined are given in the Appendix.

70% of the cohort was randomly selected to be a “development cohort” used for developing the risk prediction algorithms. The remaining 30% of the cohort was used for testing the algorithms (validation cohort). The development cohort was split further into four separate cohorts according to antipsychotic received, likewise for the validation cohort.

### Outcome/ Choice of primary measure

Our outcome was time to a new intervention for psychosis. This included: a switch to a different antipsychotic, add-on of another antipsychotic or mood stabiliser, or psychiatric hospitalisation. This composite measure was chosen to capture treatment failure and similar outcomes have been used in pragmatic randomised controlled trials in psychiatry.[25, 26] Psychiatric hospitalisations were included as indicative of increased severity of symptoms and thus treatment failure. Switches/add-ons of medications were included as indicative of the initial medication being suboptimal and thus treatment failure. Discontinuation of the medication alone was not included in the outcome as it could indicate both the medication being ineffective or the medication being effective enough for symptoms to have improved to the extent that medication is halted. The components of our outcome are well-recorded in EHRs and strongly associated with worsened quality of life.[27]

Antipsychotic medications were identified from drug issue records as records with BNF Chapter 4.2 or with SNOMED codes mapped to BNF Chapter 4.2. Psychiatric hospitalisations were identified from linked HES records as hospital APC or A&E episodes with a psychiatric primary diagnosis (ICD chapters F10-F69) and/or with a psychiatric treatment specialty and/or a record of self-harm (given as a primary or subsequent diagnosis, or as the reason for the A&E episode) and/or a symptom/ injury as a primary diagnosis (ICD T36-50, T96, Y10-34, Y87.2, R44-46, R63.6) with a psychiatric secondary diagnosis.

### Follow-up

Follow-up start (baseline) was the date of the first prescription for the antipsychotic given as first-line monotherapy. Study exit (censor date) was the earliest of: death date, date patient transferred out of the practice, date of last data collection for practice, 31 December 2019, three years after baseline. Follow-up time was time from baseline to the earliest of censor date or date of outcome.

### Potential predictors

Following sample size calculations (see Appendix) we considered an initial set of key demographic and psychiatric variables totalling 23 coefficients, to satisfy the smallest cohort sample size calculation: age at baseline, sex, ethnicity, type of SMI (schizophrenia, bipolar disorder, other psychosis), time from first SMI diagnosis to first antipsychotic prescription, prior diagnoses in the primary care records of: depression, anxiety, eating disorder, personality disorder, post-traumatic stress disorder, adverse childhood experiences, prior recorded drug misuse or alcohol misuse and the number of psychiatric hospitalisations in the previous year. We then considered a wider list of pharmacological and physical health predictors (full list in Table 1). Details of how predictors were ascertained and sample size calculations are in the Appendix.

**Table 1:**
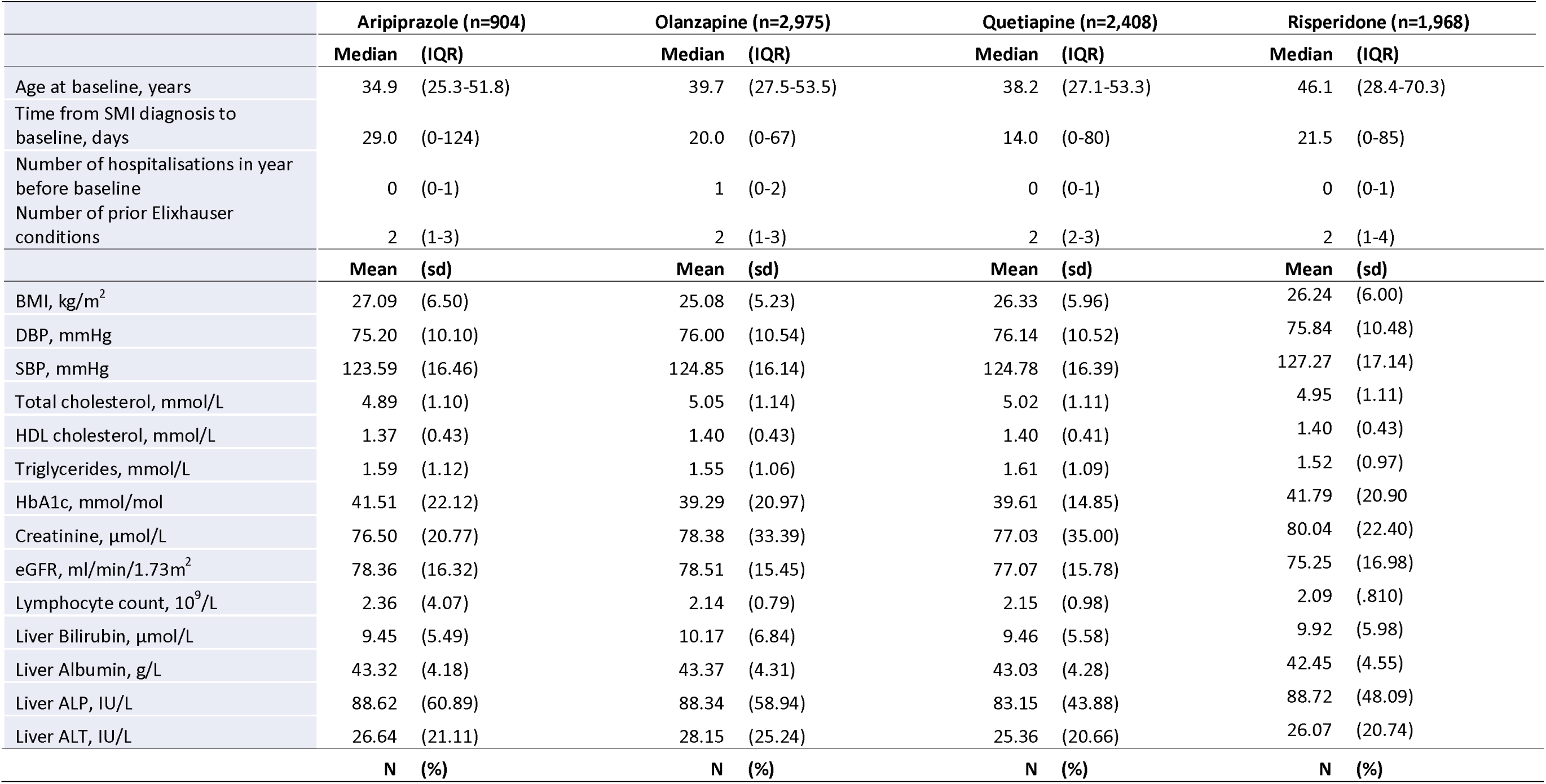
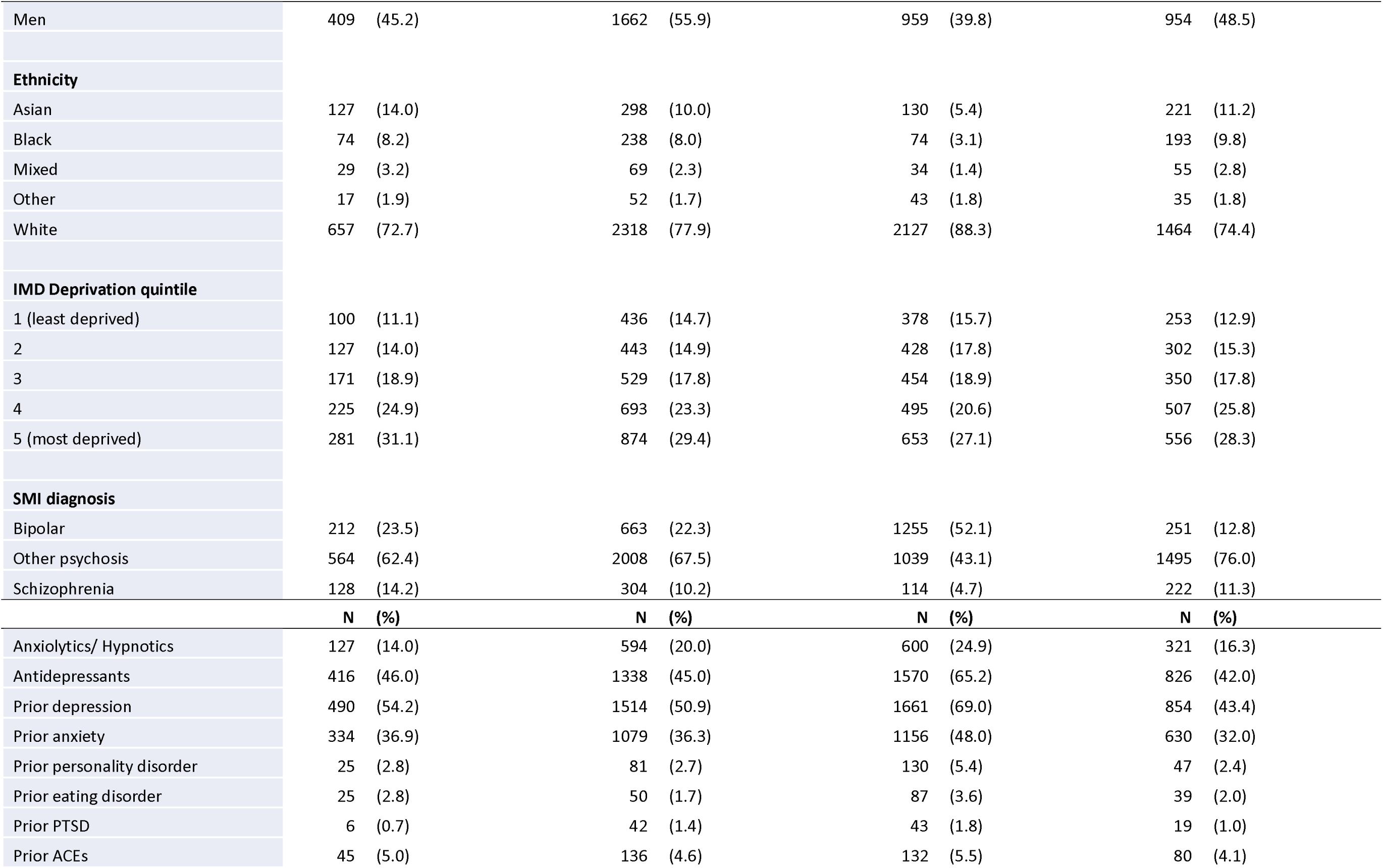
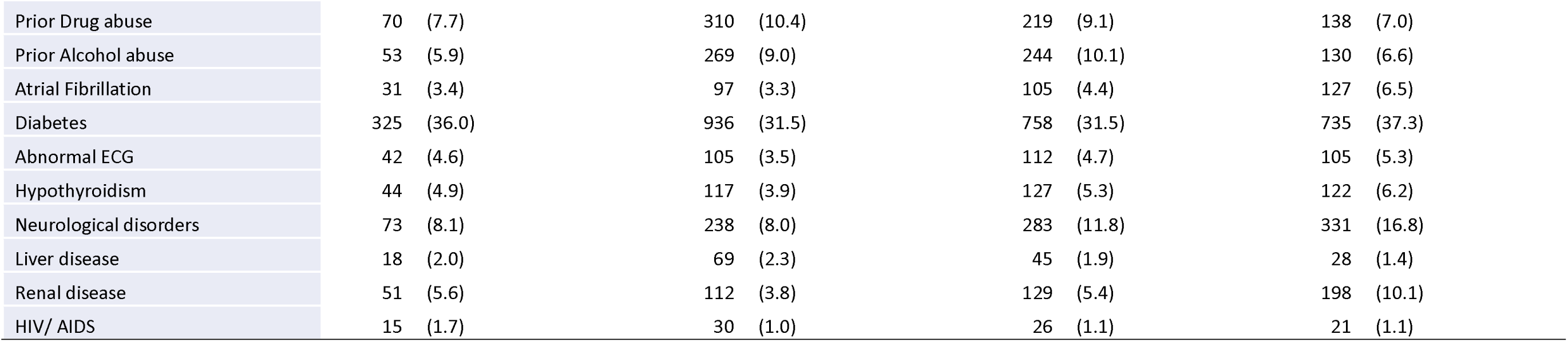
Distributions of predictor variables in the development cohorts.

|  | Aripiprazole (n=904) |  | Olanzapine (n=2,975) |  | Quetiapine (n=2,408) |  | Risperidone (n=1,968) |  |
| --- | --- | --- | --- | --- | --- | --- | --- | --- |
|  | Median | (IQR) | Median | (IQR) | Median | (IQR) | Median | (IQR) |
| Age at baseline, years | 34.9 | (25.3-51.8) | 39.7 | (27.5-53.5) | 38.2 | (27.1-53.3) | 46.1 | (28.4-70.3) |
| Time from SMI diagnosis to baseline, days | 29.0 | (0-124) | 20.0 | (0-67) | 14.0 | (0-80) | 21.5 | (0-85) |
| Number of hospitalisations in year before baseline | 0 | (0-1) | 1 | (0-2) | 0 | (0-1) | 0 | (0-1) |
| Number of prior Elixhauser conditions | 2 | (1-3) | 2 | (1-3) | 2 | (2-3) | 2 | (1-4) |
|  | Mean | (sd) | Mean | (sd) | Mean | (sd) | Mean | (sd) |
| BMI, kg/m<sup>2</sup> | 27.09 | (6.50) | 25.08 | (5.23) | 26.33 | (5.96) | 26.24 | (6.00) |
| DBP, mmHg | 75.20 | (10.10) | 76.00 | (10.54) | 76.14 | (10.52) | 75.84 | (10.48) |
| SBP, mmHg | 123.59 | (16.46) | 124.85 | (16.14) | 124.78 | (16.39) | 127.27 | (17.14) |
| Total cholesterol, mmol/L | 4.89 | (1.10) | 5.05 | (1.14) | 5.02 | (1.11) | 4.95 | (1.11) |
| HDL cholesterol, mmol/L | 1.37 | (0.43) | 1.40 | (0.43) | 1.40 | (0.41) | 1.40 | (0.43) |
| Triglycerides, mmol/L | 1.59 | (1.12) | 1.55 | (1.06) | 1.61 | (1.09) | 1.52 | (0.97) |
| HbA1c, mmol/mol | 41.51 | (22.12) | 39.29 | (20.97) | 39.61 | (14.85) | 41.79 | (20.90) |
| Creatinine, µmol/L | 76.50 | (20.77) | 78.38 | (33.39) | 77.03 | (35.00) | 80.04 | (22.40) |
| eGFR, ml/min/1.73m<sup>2</sup> | 78.36 | (16.32) | 78.51 | (15.45) | 77.07 | (15.78) | 75.25 | (16.98) |
| Lymphocyte count, 10<sup>9</sup>/L | 2.36 | (4.07) | 2.14 | (0.79) | 2.15 | (0.98) | 2.09 | (.810) |
| Liver Bilirubin, µmol/L | 9.45 | (5.49) | 10.17 | (6.84) | 9.46 | (5.58) | 9.92 | (5.98) |
| Liver Albumin, g/L | 43.32 | (4.18) | 43.37 | (4.31) | 43.03 | (4.28) | 42.45 | (4.55) |
| Liver ALP, IU/L | 88.62 | (60.89) | 88.34 | (58.94) | 83.15 | (43.88) | 88.72 | (48.09) |
| Liver ALT, IU/L | 26.64 | (21.11) | 28.15 | (25.24) | 25.36 | (20.66) | 26.07 | (20.74) |
|  | N | (%) | N | (%) | N | (%) | N | (%) |
| Men | 409 | (45.2) | 1662 | (55.9) | 959 | (39.8) | 954 | (48.5) |
| **Ethnicity** |  |  |  |  |  |  |  |  |
| Asian | 127 | (14.0) | 298 | (10.0) | 130 | (5.4) | 221 | (11.2) |
| Black | 74 | (8.2) | 238 | (8.0) | 74 | (3.1) | 193 | (9.8) |
| Mixed | 29 | (3.2) | 69 | (2.3) | 34 | (1.4) | 55 | (2.8) |
| Other | 17 | (1.9) | 52 | (1.7) | 43 | (1.8) | 35 | (1.8) |
| White | 657 | (72.7) | 2318 | (77.9) | 2127 | (88.3) | 1464 | (74.4) |
| **IMD Deprivation quintile** |  |  |  |  |  |  |  |  |
| 1 (least deprived) | 100 | (11.1) | 436 | (14.7) | 378 | (15.7) | 253 | (12.9) |
| 2 | 127 | (14.0) | 443 | (14.9) | 428 | (17.8) | 302 | (15.3) |
| 3 | 171 | (18.9) | 529 | (17.8) | 454 | (18.9) | 350 | (17.8) |
| 4 | 225 | (24.9) | 693 | (23.3) | 495 | (20.6) | 507 | (25.8) |
| 5 (most deprived) | 281 | (31.1) | 874 | (29.4) | 653 | (27.1) | 556 | (28.3) |
| **SMI diagnosis** |  |  |  |  |  |  |  |  |
| Bipolar | 212 | (23.5) | 663 | (22.3) | 1255 | (52.1) | 251 | (12.8) |
| Other psychosis | 564 | (62.4) | 2008 | (67.5) | 1039 | (43.1) | 1495 | (76.0) |
| Schizophrenia | 128 | (14.2) | 304 | (10.2) | 114 | (4.7) | 222 | (11.3) |
|  | **N** | **(%)** | **N** | **(%)** | **N** | **(%)** | **N** | **(%)** |
| Anxiolytics/ Hypnotics | 127 | (14.0) | 594 | (20.0) | 600 | (24.9) | 321 | (16.3) |
| Antidepressants | 416 | (46.0) | 1338 | (45.0) | 1570 | (65.2) | 826 | (42.0) |
| Prior depression | 490 | (54.2) | 1514 | (50.9) | 1661 | (69.0) | 854 | (43.4) |
| Prior anxiety | 334 | (36.9) | 1079 | (36.3) | 1156 | (48.0) | 630 | (32.0) |
| Prior personality disorder | 25 | (2.8) | 81 | (2.7) | 130 | (5.4) | 47 | (2.4) |
| Prior eating disorder | 25 | (2.8) | 50 | (1.7) | 87 | (3.6) | 39 | (2.0) |
| Prior PTSD | 6 | (0.7) | 42 | (1.4) | 43 | (1.8) | 19 | (1.0) |
| Prior ACEs | 45 | (5.0) | 136 | (4.6) | 132 | (5.5) | 80 | (4.1) |

|  |  |  |  |  |
| --- | --- | --- | --- | --- |
| Prior Drug abuse | 70 (7.7) | 310 (10.4) | 219 (9.1) | 138 (7.0) |
| Prior Alcohol abuse | 53 (5.9) | 269 (9.0) | 244 (10.1) | 130 (6.6) |
| Atrial Fibrillation | 31 (3.4) | 97 (3.3) | 105 (4.4) | 127 (6.5) |
| Diabetes | 325 (36.0) | 936 (31.5) | 758 (31.5) | 735 (37.3) |
| Abnormal ECG | 42 (4.6) | 105 (3.5) | 112 (4.7) | 105 (5.3) |
| Hypothyroidism | 44 (4.9) | 117 (3.9) | 127 (5.3) | 122 (6.2) |
| Neurological disorders | 73 (8.1) | 238 (8.0) | 283 (11.8) | 331 (16.8) |
| Liver disease | 18 (2.0) | 69 (2.3) | 45 (1.9) | 28 (1.4) |
| Renal disease | 51 (5.6) | 112 (3.8) | 129 (5.4) | 198 (10.1) |
| HIV/ AIDS | 15 (1.7) | 30 (1.0) | 26 (1.1) | 21 (1.1) |

### Statistical methods

#### Classical risk prediction model development

Separate prediction models for each of the four medication types were developed, first considering the key predictors totalling 23 coefficients, and then the wider list. Prediction models were derived from Cox proportional hazards multivariable regression models.

Backwards elimination using the Akaike Information Criteria was used to identify which predictors to retain in the final models. The prediction models for each medication type were then combined to form the overall treatment rule: an individual is assigned the medication for which they are computed to have the lowest overall predicted 3-year risk. Full details of model development (including imputation of missing data, use of fractional polynomials to assess form of continuous variables, categorisations of variables) are given in the Appendix, with levels of missingness reported in Appendix Table 1.

#### Model validation and comparisons

Model validity was assessed by computing Uno’s C index and Royston’s D statistic for discrimination, the calibration slope and the sensitivity and specificity at three years for different risk thresholds. Each medication risk prediction model was validated in the separate validation cohort for that medication. Details of the model validations are given in the Appendix.

The utility of the overall treatment rule was investigated by comparing the predicted 3-year risk if all individuals are assigned the medication dictated by the risk prediction models (medication which corresponds to lowest predicted risk for the individual) with i) the outcome under the observed medications; ii) the predicted outcome if all people were to receive the same one medication (for each of the four medications in turn); iii) the predicted outcome if all patients were randomly allocated one of the four medications. Absolute risk reductions, and corresponding numbers needed to treat, comparing the risk prediction model medication assignment with each of the other scenarios were computed. In addition, individuals were grouped into two categories: those whose recorded medication was the same as predicted and those whose recorded medication differed from that predicted.

Then the outcome was compared between the two groups.

#### Machine learning model development

In addition to the classical Cox regression models, several machine learning models (adapted for survival data) were considered: random forest, extreme gradient boosted model, linear and kernel support vector machines, deep learning neural networks and a Super Learner ensemble, combining all the models together. Details of the machine learning models are given in the Appendix.

## Results

### Cohort characteristics

A total of 11,811 individuals with SMI were identified for inclusion in the analysis, as having received aripiprazole, olanzapine, quetiapine or risperidone as a first-line monotherapy, between 2007-2016. Of these, 8,225 were included in the development cohorts: 904 (11%) having received aripiprazole forming the aripiprazole development cohort, and 2,975 (36%), 2,408 (29%), and 1,968 (24%) in the olanzapine, quetiapine and risperidone cohorts respectively. The remaining 3,556 individuals constituted the validation cohorts, including 397 (11%) individuals having received aripiprazole, and 1,306 (37%), 1,027 (29%) and 826 (23%) having received olanzapine, quetiapine and risperidone respectively. Distributions of predictors in the development cohorts according to medication are shown in table 1.

Predictor distributions in the validation cohorts were broadly similar (Appendix Table 2).

### Treatment outcomes

In the aripiprazole development cohort, outcome events occurred within three years in 456 individuals over 1,487 person-years of follow-up, corresponding to an incidence rate of 306.8 per 1000 person-years at risk (PYAR), 95% confidence interval (CI) 279.9 to 336.3 (Table 2). Among the 456 events, 261 (57.2%) were hospitalisations, 150 (32.9%) were medication switches and 45 (9.9%) were medication add-ons (Appendix table 3). Incidence rates in the olanzapine, quetiapine and risperidone cohorts were: 351.5 (95% CI 335.0 to 368.9), 356.7 (95% CI 338.0 to 376.5) and 332.9 (313.2 to 353.9) respectively. Similar proportions of events were hospitalisations in these groups as for aripiprazole. The five most common medication switches and add-ons in each medication cohort are shown in Appendix table 4.

**Table 2:** Incidence rates of treatment failure (hospitalisation and/or medication changes/ add-ons) in the development and validation cohorts Development cohorts.

|  | Outcome | Person years | Incidence rates per 1000 PYAR (95% CI) |
| --- | --- | --- | --- |
| Aripiprazole | 456 | 1487 | 306.8 (279.9, 336.3) |
| Olanzapine | 1654 | 4705 | 351.5 (335, 368.9) |
| Quetiapine | 1319 | 3698 | 356.7 (338, 376.5) |
| Risperidone | 1027 | 3085 | 332.9 (313.2, 353.9) |

Validation cohorts
|  | Outcome | Person years | Incidence rates per 1000 PYAR (95% CI) |
| --- | --- | --- | --- |
| Aripiprazole | 216 | 654 | 330.4 (289.2, 377.6) |
| Olanzapine | 691 | 2175 | 317.8 (294.9, 342.4) |
| Quetiapine | 579 | 1529 | 378.8 (349.2, 410.9) |
| Risperidone | 430 | 1292 | 332.9 (302.8, 365.8) |

Incidence rates of outcome events in the validation cohorts were: aripiprazole 330.4 (95% CI 289.2 to 377.6), olanzapine 317.8 (95% CI 294.9 to 342.4), quetiapine 378.8 (95% CI 349.2 to 410.9) and risperidone 332.9 (95% CI 302.8 to 365.8), per 1000 PYAR.

### Cox regression models development

The final risk prediction models, after backwards elimination for each medication starting from the full list of predictors are shown in table 3. Key retained predictors included age, sex, ethnicity, time from SMI diagnosis to first prescription, SMI diagnosis, prior depression, anxiety, alcohol misuse and number of prior psychiatric hospitalisations. The final risk prediction models after backwards elimination, developed starting instead from a limited number of predictors totalling 23 coefficients are shown in Appendix table 5.

**Table 3:** Classical statistical prediction models after backwards elimination – starting from full list of predictors.

| Predictor | Coefficient95% CI | HR95% CI | p |
| --- | --- | --- | --- |
| Age | -0.003(-0.010, 0.003) | 0.997(0.99, 1.003) | 0.3 |
| Age squared | 0.000(0.000, 0.001) | 1(1, 1.001) | 0.006 |
| ALT liver test | -0.008(-0.015, -0.001) | 0.992(0.985, 0.999) | 0.04 |
| ALT squared | 0.000(0.000, 0.000) | 1.000(1.000, 1.000) | 0.1 |
| Lymphocyte count | -0.087(-0.217, 0.044) | 0.917(0.805, 1.045) | 0.2 |
| Lymphocyte count squared | 0.001(0.000, 0.002) | 1.001(1.000, 1.002) | 0.1 |
| Ethnicity |  |  |  |
| Asian | -0.142(-0.433, 0.150) | 0.868(0.648, 1.162) |  |
| Black | -0.054(-0.416, 0.307) | 0.947(0.660, 1.360) |  |
| Mixed | 0.396(-0.156, 0.947) | 1.485(0.856, 2.578) |  |
| Other | -1.147(-2.048, -0.247) | 0.318(0.129, 0.782) |  |
| White | 0 | 1 | 0.05 |
| SMI Diagnosis |  |  |  |
| Bipolar | 0.374(0.156, 0.592) | 1.453(1.169, 1.807) |  |
| Other psychosis | 0 | 1 |  |
| Schizophrenia | -0.137(-0.427, 0.153) | 0.872(0.652, 1.166) | <0.001 |
| Prior personality disorder | 0.408(-0.108, 0.924) | 1.504(0.898, 2.519) | 0.1 |
| Prior hospitalisations |  |  |  |
| 0 | 0 | 1 |  |
| 1 | 0.178(-0.053, 0.409) | 1.195(0.949, 1.505) |  |
| 2 | 0.321(0.044, 0.599) | 1.379(1.045, 1.820) |  |
| 3 | -0.224(-0.712, 0.263) | 0.799(0.491, 1.301) |  |
| 4+ | 0.875(0.472, 1.278) | 2.399(1.603, 3.590) | <0.001 |
| Prior antidepressant use | 0.285(0.090, 0.480) | 1.330(1.094, 1.616) | 0.004 |
| Triglycerides | 0.101(0.006, 0.195) | 1.106(1.006, 1.216) | 0.04 |
| HDL | -0.169(-0.433, 0.096) | 0.845(0.648, 1.101) | 0.2 |
| SBP | 0.007(-0.001, 0.014) | 1.007(0.999, 1.014) | 0.08 |
| DBP | -0.010(-0.021, 0.001) | 0.990(0.979, 1.001) | 0.07 |
| Neurological disorders | -0.288(-0.667, 0.091) | 0.750(0.513, 1.095) | 0.1 |

Olanzapine
| Predictor | Coefficient95% CI | HR95% CI | p |
| --- | --- | --- | --- |
| Age | -0.011(-0.014, -0.007) | 0.989(0.986, 0.993) | <0.001 |
| Age squared | 0.000(0.000, 0.000) | 1.000(1.000, 1.000) | <0.001 |
| Women | 0.143(0.041, 0.246) | 1.154(1.042, 1.279) | 0.006 |
| SMI Diagnosis |  |  |  |
| Bipolar | 0.189(0.073, 0.305) | 1.208(1.076, 1.356) |  |
| Other psychosis | 0 | 1 |  |
| Schizophrenia | 0.184(0.025, 0.343) | 1.203(1.026, 1.410) | 0.001 |
| Prior depression | 0.124(0.010, 0.239) | 1.133(1.010, 1.270) | 0.03 |
| Prior anxiety | 0.094(-0.015, 0.203) | 1.098(0.985, 1.225) | 0.09 |
| Prior alcohol abuse | 0.176(0.013, 0.340) | 1.193(1.013, 1.405) | 0.03 |
| Prior hospitalisations |  |  |  |
| 0 | 0 | 1 |  |
| 1 | 0.173(0.057, 0.289) | 1.189(1.058, 1.335) |  |
| 2 | 0.202(0.055, 0.348) | 1.224(1.057, 1.417) |  |
| 3 | 0.293(0.102, 0.484) | 1.340(1.107, 1.622) |  |
| 4+ | 0.454(0.235, 0.673) | 1.575(1.266, 1.960) | <0.001 |
| Prior hypnotic/anxiolytic use | 0.109(-0.011, 0.229) | 1.115(0.989, 1.257) | 0.08 |
| Prior antidepressant use | 0.227(0.115, 0.339) | 1.255(1.122, 1.404) | <0.001 |
| BMI | 0.011(0.001, 0.020) | 1.011(1.001, 1.020) | 0.03 |
| ALP Liver test | 0.000(0.000, 0.001) | 1.000(1.000, 1.001) | 0.4 |
| Albumin liver test |  |  |  |
| <35 | 0.487(0.219, 0.755) | 1.627(1.245, 2.127) |  |
| 35-50 | 0 | 1 |  |
| >50 | -0.072(-0.294, 0.150) | 0.930(0.745, 1.161) | 0.001 |
| eGFR | -0.004(-0.007, 0.000) | 0.996(0.993, 1.000) | 0.04 |
| Triglycerides | -0.075(-0.142, -0.008) | 0.928(0.868, 0.992) | 0.03 |
| Total cholesterol | -0.047(-0.097, 0.003) | 0.954(0.907, 1.003) | 0.07 |
| Abnormal ECG | 0.260(0.003, 0.517) | 1.297(1.003, 1.677) | 0.05 |

Quetiapine
| Predictor | Coefficient95% CI | HR95% CI | p |
| --- | --- | --- | --- |
| Age | -0.004(-0.008, 0.000) | 0.996(0.992, 1.000) | 0.03 |
| Age squared | 0.000(0.000, 0.000) | 1.000(1.000, 1.000) | 0.03 |
| Time from diagnosis | -0.001(-0.001, 0.000) | 0.999(0.999, 1.000) | 0.005 |
| Time squared | 0.000(0.000, 0.000) | 1.000(1.000, 1.000) | 0.001 |
| Women | 0.118(-0.001, 0.236) | 1.125(0.999, 1.266) | 0.05 |
| Ethnicity |  |  |  |
| Asian | -0.144(-0.404, 0.116) | 0.866(0.667, 1.123) |  |
| Black | 0.337(0.043, 0.632) | 1.401(1.043, 1.881) |  |
| Mixed | -0.341(-0.823, 0.141) | 0.711(0.439, 1.152) |  |
| Other | 0.021(-0.395, 0.437) | 1.021(0.674, 1.548) |  |
| White | 0 | 1 | 0.1 |
| Lymphocyte count | -0.075(-0.155, 0.005) | 0.927(0.856, 1.005) | 0.06 |
| Prior anxiety | 0.191(0.077, 0.305) | 1.211(1.080, 1.357) | 0.001 |
| Prior alcohol abuse | 0.194(0.021, 0.367) | 1.214(1.021, 1.444) | 0.03 |
| Prior hospitalisations |  |  |  |
| 0 | 0 | 1 |  |
| 1 | 0.065(-0.077, 0.207) | 1.067(0.926, 1.229) |  |
| 2 | 0.385(0.201, 0.570) | 1.470(1.222, 1.768) |  |
| 3 | 0.495(0.229, 0.761) | 1.640(1.257, 2.140) |  |
| 4+ | 0.974(0.711, 1.238) | 2.650(2.036, 3.448) | <0.001 |
| Prior hypnotic/anxiolytic use | 0.091(-0.033, 0.214) | 1.095(0.968, 1.239) | 0.1 |
| Prior antidepressant use | 0.256(0.129, 0.383) | 1.292(1.137, 1.467) | <0.001 |
| Total cholesterol | 0.047(-0.003, 0.098) | 1.048(0.997, 1.103) | 0.07 |
| HDL cholesterol | -0.167(-0.329, -0.004) | 0.846(0.719, 0.996) | 0.04 |
| SBP | 0.006(0.001, 0.010) | 1.006(1.001, 1.010) | 0.01 |
| DBP | -0.009(-0.016, -0.003) | 0.991(0.984, 0.997) | 0.005 |
| Liver disease | 0.401(0.048, 0.755) | 1.494(1.049, 2.127) | 0.03 |

Risperidone
| Predictor | Coefficient 95% CI | HR 95% CI | p |
| --- | --- | --- | --- |
| Age | -0.006 (-0.010, -0.003) | 0.994 (0.990, 0.997) | <0.001 |
| Age squared | 0.000 (0.000, 0.000) | 1.000 (1.000, 1.000) | 0.03 |
| Time from diagnosis | -0.001 (-0.002, 0.000) | 0.999 (0.998, 1.000) | 0.001 |
| Time squared | 0.000 (0.000, 0.000) | 1.000 (1.000, 1.000) | 0.01 |
| Women | 0.136 (0.001, 0.271) | 1.146 (1.001, 1.311) | 0.05 |
| **SMI Diagnosis** |  |  |  |
| Bipolar | 0.204 (0.026, 0.382) | 1.226 (1.026, 1.465) |  |
| Other psychosis | 0 | 1 |  |
| Schizophrenia | 0.174 (-0.017, 0.365) | 1.190 (0.984, 1.440) | 0.03 |
| Prior depression | 0.177 (0.031, 0.323) | 1.194 (1.032, 1.382) | 0.02 |
| Prior anxiety | 0.117 (-0.024, 0.257) | 1.124 (0.976, 1.293) | 0.1 |
| Prior eating disorder | 0.300 (-0.089, 0.690) | 1.350 (0.915, 1.994) | 0.1 |
| Prior alcohol abuse | 0.293 (0.061, 0.526) | 1.341 (1.063, 1.692) | 0.01 |
| **Prior hospitalisations** |  |  |  |
| 0 | 0 | 1 |  |
| 1 | 0.239 (0.084, 0.393) | 1.269 (1.087, 1.482) |  |
| 2 | 0.430 (0.228, 0.633) | 1.538 (1.256, 1.882) |  |
| 3 | 0.489 (0.227, 0.751) | 1.631 (1.255, 2.118) |  |
| 4+ | 0.619 (0.286, 0.952) | 1.857 (1.331, 2.591) | <0.001 |
| Prior antidepressant use | 0.162 (0.016, 0.307) | 1.176 (1.017, 1.360) | 0.03 |
| BMI | -0.009 (-0.021, 0.003) | 0.991 (0.980, 1.003) | 0.1 |
| Bilirubin | -0.012 (-0.023, 0.000) | 0.988 (0.977, 1.000) | 0.05 |
| HbA1c | 0.004 (0.001, 0.008) | 1.004 (1.001, 1.008) | 0.03 |
| Triglyceride | -0.039 (-0.116, 0.037) | 0.962 (0.891, 1.038) | 0.3 |

### Cox regression models validation and decision rule evaluation

Discrimination and calibration statistics and sensitivities and specificities at different risk thresholds for the risk prediction models applied to the validation cohorts are presented in Appendix table 6. Medications predicted by the selection tool based on these Cox models for each individual in the validation cohorts are tabulated against the actual first-line treatment received in table 4. Models derived from the full list of predictors were used for the selection tool, but results were similar using the models derived from predictors totalling 23 coefficients. More than half of those originally prescribed aripiprazole as first-line treatment were also assigned aripiprazole by the personalised selection tool (58%).

**Table 4:** Medications assigned by decision rule versus medications actually prescribed in the validation cohort.

|  |  | Predicted best drug (using models with all predictors) |  |  |  |  |
| --- | --- | --- | --- | --- | --- | --- |
|  |  | Aripiprazole | Olanzapine | Quetiapine | Risperidone | Total |
| Original drug | Aripiprazole | 229 (58%) | 61 (15%) | 63 (16%) | 44 (11%) | 397 |
|  | Olanzapine | 781 (60%) | 233 (18%) | 178 (14%) | 114 (9%) | 1,306 |
|  | Quetiapine | 477 (46%) | 135 (13%) | 297 (29%) | 118 (11%) | 1,027 |
|  | Risperidone | 451 (55%) | 130 (16%) | 110 (13%) | 135 (16%) | 826 |

However, only 18%, 29% and 16% of those prescribed olanzapine, quetiapine and risperidone were assigned the same medication by the decision rule. In all 2,662 (75%) of the those in the validation cohort were assigned a different medication by the selection tool. The mean predicted 3-year risk using our selection tool was 6.3% lower than the observed risk (absolute risk reduction 6.3% (95% CI 4.0% to 8.5%)), corresponding to a number needed to treat of 16 (95% CI 12 to 25). The mean predicted 3-year risk using the treatment rule was between 4% and 13% lower than in the scenario where all individuals were assigned any one medication, and 10% lower than in the scenario of random allocation (Table 5). Further, as a separate evaluation, among individuals in the development cohort who were assigned the same medication by the treatment rule as the medication they were originally prescribed, the observed 3-year risk was 58.8%, while among those for whom a different medication was assigned by the treatment rule, the observed 3-year risk was 55.0% (relative risk of 1.07).

**Table 5:** Predicted 3-year risk of treatment failure using decision rule and other treatment rules versus observed 3-year risk of treatment failure in validation cohort.

| Treatment rule | Predicted 3-year risk | ARR using predicted best drug rule (95% CI)* | NNT (=1/ARR) (95%CI)** |
| --- | --- | --- | --- |
| **Predicted best drug** | **51.5%** |  |  |
| All Aripiprazole | 56.2% | 4.4 (2.1, 6.7) | 23 (15, 48) |
| All Olanzapine | 63.2% | 12.0 (9.7, 14.2) | 8 (7,10) |
| All Quetiapine | 62.7% | 11.1 (8.9, 13.5) | 9 (7, 11) |
| All Risperidone | 64.8% | 13.3 (11.0, 15.6) | 8 (6, 9) |
| Random selection | 61.5% | 10.2 (7.9, 12.5) | 10 (8, 13) |
| **Observed risk** | **57.8%** | **6.3 (4.0, 5.8)** | **16 (12, 25)** |

\*ARR = Absolute Risk Reduction comparing predicted best drug rule against the other treatment rules (rows 2-6) and against the observed risk (row 7)
\*\*NNT = Number Needed to Treat Reduction comparing predicted best drug rule against the other treatment rules (rows 2-6) and against the observed risk (row 7)

### Machine learning models development and validation

Discrimination statistics from application of the machine learning models to the validation cohorts are shown in Appendix table 7. Of the machine learning models, the extreme gradient boosted models appeared to have consistently stronger performance across all four medication groups, although differences in C statistics between the different models were marginal. The machine learning models, including the Super Learner ensemble, did not materially outperform the classical statistical survival models.

## Discussion

We successfully developed a treatment selection tool to identify the optimum of four medications: aripiprazole, olanzapine, quetiapine and risperidone for use as first-line monotherapy in SMI. The decision rule consisted of applying risk prediction models specific to each medication, encompassing a range of clinical and demographic predictors, to an individual, and then selecting the medication for which the corresponding risk prediction model predicted the lowest 3-year risk of treatment failure (hospitalisation or a change in medication). The tool was applied to a separate validation cohort of individuals who had received one of these medications. We found that had all individuals received the medication indicated by the decision rule, 75% of individuals would have been prescribed a different medication from that actually received. The 3-year risk of treatment failure in the cohort could have been 6% lower than that observed, corresponding to a number needed to treat of 16, and 10% lower than had medications been randomly assigned.

To our knowledge, this is the first study to develop a treatment decision rule for all incident SMI (schizophrenia, bipolar disorder and other psychosis) and in populations outside of Asia. A previous study in Taiwan which developed a treatment rule using machine learning for first episode schizophrenia, found a similar difference of 7% between predicted treatment success using the treatment rule, and that observed (52% versus 45%).[21] In agreement with the present study, they also found that aripiprazole was the most frequently recommended medication by the treatment rule, lending further support to our findings.

Another previous study developed a treatment rule for children and adolescents with schizophrenia in South Korea.[20] That study reported a ratio of treatment continuation (study outcome) of those receiving the medication assigned by the treatment rule relative to those not receiving the assigned medication of 1.2 to 1.5. This is higher than the 1.07 relative risk of treatment failure comparing these two groups in our study. However, the outcome in that study is not directly comparable to our study as it included stopping the medication, as opposed to just switching or adding to the medication. As outlined in the Methods, this is problematic as stopping the medication could indicate treatment success as well as treatment failure. Interestingly, the study in South Korea also compared classical statistical (logistic) regression models with machine learning models and in agreement with our study, found machine learning models did not materially improve performance. The authors attribute the lack of superiority of machine learning models in part to the use of clinically meaningful predictors such that machine learning models had little to add, a finding supported by previous studies using structured data with clinical meaningful predictors[28, 29]. This could also be the case in our study. Further, we carefully examined all continuous predictors to assess the appropriate form/transformation in the statistical models, thus the added benefit of machine learning models in capturing non-linear and higher order effects may be limited.

Strengths of the study include the wide range of well-defined clinical and demographic predictors available for inclusion in the models, and which reflect the information available to clinicians in practice, thus supporting potential implementation. In addition, the study included a large sample of individuals with SMI receiving first-line monotherapy of one of the four medications of interest, thus the statistical models were powered to include the large number of predictors available. Previous studies have shown people with SMI in UK primary care databases to be broadly representative of the UK SMI population.[24] A study limitation is the lack of information on medication dose: the treatment decision rule thus only assigns the type of medication and not the dose. The modelling implicitly assumes that the individual is receiving the optimum dose, but treatment failure could be due to the dose being incorrect, rather that the type of medication being suboptimal. However, in the course of treating a patient, the dose is likely to be adjusted and optimised before resorting to changing the medication, thus dose should not unduly influence the results. Secondly, medication adherence is unknown, although we only included people with at least one repeat prescription to indicate the individual adhering to the medication (see Appendix). A third limitation is the nature of the CPRD, comprising a routinely-collected snapshot of an individual’s primary care records, for the period during which the individual is included in the CPRD, which means that historical patient records prior to an individual registering with a general practice that contributes data to CPRD may not be captured. This means that historical antipsychotic prescriptions may be missed such that the first-line treatment as defined here is not the true first antipsychotic medication. However, we only included individuals in the study, for whom the first recorded antipsychotic medication use occurred at least one year after the patient entered the study (Appendix). This helped to minimise the possibility of prior uncaptured antipsychotic medication use. A further limitation is that although a large number of relevant predictors were included, some information was not available which may have equally limited the predictive power of both the machine learning models and the statistical models, specifically genetic factors, and information on severity and clinical presentation of SMI. Future research could explore the influence of these additional factors, as well as potential modifications to try to improve the machine learning models. Further research will also consider the combined impact of the treatment rule on treatment failure (effectiveness outcome) and adverse outcomes (weight gain, hypertension, hypercholesterolemia and diabetes), and the application of the treatment rule in different populations. Future research should also consider cost-effectiveness of the treatment rule when used in routine clinical practice.

In conclusion, the results of this study suggest that use of a personalised treatment selection tool in SMI to assign first-line medications could help to reduce the risk of treatment failure in severe mental illness. Our treatment rule was developed using classical, clearly defined, easily characterised and interpreted statistical models, using a range of predictors available in UK national health data. Although developed using primary care data, the models should be applicable in other health services such as secondary care psychiatric services, where antipsychotics tend to be first prescribed. This means the tool could feasibly be implemented in UK practice. Further work is necessary to prospectively validate the models. As we found little improvement in performance using machine learning, integration of our simple statistical models into clinical workflows would be relatively straightforward. Low risk prediction decisions, such as selection of treatment from a number of approved options, could improve outcomes for people with SMI.

## Funding

This research is funded by UKRI grant MR/V023373/1, the University College London Hospitals NIHR Biomedical Research Centre and the NIHR North Thames Applied Research Collaboration (JFH).

NL is supported by a HDR UK personal fellowship (Big Data for Complex Disease driver programme: HDR-23012). This work is affiliated to HDR UK which is funded by the Medical Research Council (UKRI), the National Institute for Health Research, the British Heart Foundation, Cancer Research UK, the Economic and Social Research Council (UKRI), the Engineering and Physical Sciences Research Council (UKRI), Health and Care Research Wales, Chief Scientist Office of the Scottish Government Health and Social Care Directorates, and Health and Social Care Research and Development Division (Public Health Agency, Northern Ireland). NL is additionally supported by the UK Research and Innovation grant MR/W014386/1 and MR/V023373/1 and by the NIHR University College London Hospitals Biomedical Research Centre, and NIHR North Thames Applied Research Collaboration.

ARB is funded by the Wellcome Trust through a PhD Fellowship in Mental Health Science (218497/Z/19/Z). This research was funded in whole or in part by the Wellcome Trust. For the purpose of Open Access, the author has applied a CC BY public copyright licence to any Author Accepted Manuscript (AAM) version arising from this submission.

## Data Availability

Study data were accessed via Clinical Practice Research Datalink (CPRD). Authors cannot share the data directly. However data can be accessed directly from CPRD following approval and licensing.

## Notes

### Competing Interest Statement

The authors have declared no competing interest.

### Author Declarations

The East Midlands - Derby Research Ethics Committee gave ethical approval for the Clinical Practice Research Datalink (CPRD) database used for this work (reference: 21/EM/0265). The Independent Scientific Advisory Committee of CPRD gave ethical approval for this work (protocol number: #21_000729).

