## Supplementary material for "Development and validation of a personalised antipsychotic selection tool for first-line treatment in severe mental illness"

**Table of contents**

1-3	Methods: Additional details

5-6	Appendix table 1: Levels of missingness of incomplete predictor variables

7-9	Appendix table 2: Distributions of predictor variables in the validation cohorts

9	Appendix table 3: Types of outcomes in each development and validation cohort

10	Appendix table 4: The five most common medication changes and add-ons in each development cohort

11-15	Appendix table 5: Classical statistical prediction models after backwards elimination – starting from predictors totalling 23 coefficients

16	Appendix table 7: Discrimination (C) statistics for machine learning risk prediction models compared with classical statistical risk prediction models

17	References for Appendix

**Methods: additional details**

**Additional details of Cohort**

As outlined in the Methods, the cohort comprised individuals with incident SMI between January 2007 and December 2016, who commenced on one of four antipsychotics as a first-line monotherapy treatment: aripiprazole, quetiapine, olanzapine, and risperidone. To be considered to have had first-line treatment with one of the four antipsychotics, the antipsychotic had to be the first antipsychotic prescription in the patient records, and given in isolation as a monotherapy, without other antipsychotics at the same time. Thus individuals prescribed any other antipsychotics or lithium, lamotrigine or valproate before one of the four listed were excluded. Individuals prescribed one of the four main drugs in combination with another antipsychotic as first-line treatment were also excluded. In addition to the inclusion and exclusion criteria outlined in the Methods, patients receiving injectable versions of the four antipsychotics, given on a repeated basis, were excluded as these are unlikely to be first-line treatments. However, patients prescribed a one-off injectable antipsychotic prior to one of the four drugs were retained. To ensure first-line treatment, only patients with incident SMI were included, defined as having first record of SMI prior to, or within six months after, the first prescription (the six month window after prescription was to allow for delays in transfer of diagnoses to GP records). Following exploratory work on timings of diagnoses and prescriptions, we further excluded patients whose SMI diagnosis occurred more than four years prior to the first prescription as these were largely historical records preceding registration at the GP practice and thus we could not guarantee the patient did not receive antipsychotics before the first record in CPRD. Only patients with a second follow-on prescription within 3 months of the first for the given drug were included, to confirm as far as possible that the first prescription was used, warranting a reissue. Individuals who exited the study (died or transferred out of the practice or end of data collection) before 3 months were excluded to avoid immortal time bias.

Individuals with no available data in CPRD 12 months before the date of initiation of antipsychotic treatments were excluded since we would not know when they initiated treatment. This also ensured sufficient time to capture baseline characteristics and predictors. We further excluded individuals with no CPRD data 12 months before the incident SMI record (if earlier than the date of first antipsychotic). Finally, people for whom linkage to HES was not available were excluded i.e. non-English practices (13 practices, 91 people) plus a small number of English practices (8 practices, 60 people), plus individuals within the 1,405 linked English practices who were not eligible (291 people).

**Sample size calculations**

Sample size calculations indicated risk prediction models were powered to include up to 23 coefficients for the smallest medication development cohort (aripiprazole) and over 50 in the other development cohorts (olanzapine, quetiapine and risperidone), based on 20 events required per coefficient to achieve a satisfactory level of model calibration[1] and 456 events in the aripiprazole development cohort, and 1,654, 1,319, and 1,027 events in the olanzapine, quetiapine, and risperidone cohorts.

For model validation, studies suggest needing at least 100 events, and ideally 200 events, in a validation cohort [2]. This was achieved in all four validation cohorts (216, 691, 579, and 430 events in the aripiprazole, olanzapine, quetiapine, and risperidone validation cohorts respectively).

**Additional details of Potential predictors**

As outlined in the Methods, to satisfy the aripiprazole sample size calculation, we first considered key demographic and psychiatric variables totalling 23 coefficients: age at baseline, sex, ethnicity, type of SMI (schizophrenia, bipolar disorder, other psychosis), time from first SMI diagnosis to first antipsychotic prescription, prior diagnoses in the primary care records of: depression, anxiety, eating disorder, personality disorder, post-traumatic stress disorder, adverse childhood experiences, prior recorded drug misuse or alcohol misuse and the number of psychiatric hospitalisations in the previous year.

Since the other medication cohorts were large enough to allow for a greater number of coefficients, we also considered a wider range of pharmacological and physical health predictors: all predictors above plus receipt of other psychotropic medications (antidepressants, anxiolytics and hypnotics) in the previous year, blood tests (nearest prior to baseline) for HbA1c, creatinine, eGFR, triglycerides, total cholesterol, HDL cholesterol, lymphocyte count, liver function: ALT, ALP, albumin and bilirubin, nearest prior to baseline BMI and blood pressure, Index of Multiple Deprivation (IMD, based on patient postcode), prior diagnoses in primary care records of: atrial fibrillation, diabetes, abnormal ECG, hypothyroidism, neurological disorders (including epilepsy, multiple sclerosis, Parkinson disease, and seizures), liver disease, renal disease, HIV/AIDS, and the total number of conditions in the Elixhauser comorbidity index diagnosed prior to baseline [3]. Choice of predictors was based on variables which might plausibly be related to the risk of the outcome and variables which were available in the CPRD records.

Ethnicity was initially ascertained from CPRD records, and where missing, was determined from the linked HES records. Diagnoses were identified using code lists developed for each condition, and prior medications from drug issue records with relevant BNF chapters (4.1 for anxiolytics and hypnotics, 4.3 for antidepressants). Adverse childhood experiences was identified from records indicating contact with social services, child protection procedures, childhood maltreatment (physical or emotional abuse or neglect, as defined in the Centers for Disease Control and Prevention short adverse childhood experiences tool [4]) or household dysfunction (such as parental substance misuse). Prior psychiatric hospitalisations were identified from HES in the same way as the incident outcome hospitalisations as hospital APC or A&E episodes with a psychiatric primary diagnosis (ICD chapters F10-F69) and/or with a psychiatric treatment specialty and/or a record of self-harm (given as a primary or subsequent diagnosis, or as the reason for the A&E episode) and/or a symptom/ injury as a primary diagnosis (ICD T36-50, T96, Y10-34, Y87.2, R44-46, R63.6) with a psychiatric secondary diagnosis. Blood test results, BMI and blood pressure measurements were extracted from CPRD records, converted to consistent, standard units where needed and outliers were removed before the closest record prior to baseline was identified. Outliers were identified as values outside previously defined ranges from resources including UK biobank[5] as well as within-person outliers using published methods for EHR data [6]. The Elixhauser comorbidity index includes asthma, COPD, cardiac arrhythmia, congestive heart failure, myocardial infarction, cerebrovascular disease, neurological disorders, cancer, diabetes (type 1 or 2), hypothyroidism, liver disease, renal disease, peptic ulcers, rheumatic and collagen disease, paresis or paralysis, HIV/AIDS, hypertension, peripheral vascular disease, pulmonary circulation disorders, valvular disease, deficiency anaemia, blood loss anaemia, coagulopathy, and fluid or electrolyte disorders.

**Statistical Methods**

***Classical risk prediction model development***

As outlined in the Methods, separate prediction models were developed for each of the four medication types: aripiprazole, olanzapine, quetiapine and risperidone, derived from Cox proportional hazards regression models. Developing separate models for each medication as opposed to a single combined model, allowed the relationship between all the predictors and the outcome to differ between medication types, thus potentially offering better model fit. Models were developed first by considering the key predictors totalling 23 coefficients and then the full list of coefficients above.

In the Cox proportional hazards models, sex, all diagnoses and all medications were included as indicator variables. Ethnicity was categorised as: Asian, Black, Mixed, Other, White. Type of SMI was categorised as: schizophrenia, bipolar disorder and other psychosis. IMD was available (and included) as quintiles, based on the CPRD population as a whole. After examining frequencies and outcome rates for count variables (number of prior psychiatric hospitalisations and number of prior Elixhauser conditions), categories with similar outcome rates were groups together, so that number of prior psychiatric hospitalisations was categorised as 0,1,2,3 or more and number of prior Elixhauser conditions was categorised as 0,1,2,3,4,5,6, or more.

Fractional polynomial regression modelling and graphs were used to assess appropriate forms for the continuous variables in the models (namely age, time from first diagnosis to first prescription, ALT, ALP, albumin, bilirubin, lymphocyte count, eGFR, creatinine, HbA1c, triglyceride, total cholesterol, HDL cholesterol and blood pressure)[7]. Based on fractional polynomial regression modelling and graphical assessments, in the aripiprazole model, squared terms were included for age, ALT and lymphocyte count; in the olanzapine model, squared terms were included for age and lymphocyte count; in the quetiapine model, squared terms were included for age and time from first diagnosis to first prescription; and in the risperidone model, squared terms were included for time from first diagnosis to first prescription. Since these assessments showed isolated high risk for high and low albumin levels, in line with recognised risk patterns, standard classifications were used to categorise albumin (as <35g/L (low), 35-50g/L (normal) and >50g/L (high)). In exploring the data, we found eGFR values were concentrated around standard categorisation cut-offs, rather than distributed as a continuous spread, suggesting that eGFR tends to be inputted into records as categories rather than the actual measured level. Thus eGFR was categorised using standard cut-offs: (<15ml/min (kidney failure), 15-60ml/min (kidney disease), 60-90ml/min (early stage kidney disease), ≥90ml/min (normal)).

Diagnoses variable (including type of SMI and number of Elixhauser conditions), medication variables, and number of prior psychiatric hospitalisations, were assumed complete (so the absence of the diagnosis record or medication prescription record was taken to mean the absence of the condition or medication). The very small number of people missing sex (2 people, 0.01% in the development and validation cohorts combined) or IMD (17 people, 0.1% of the combined cohorts) were excluded. The small number of people missing ethnicity (65, 0.5% of the combined cohorts) were recoded as “Other”. Multiple imputation [8, 9]was used to impute missing data in HbA1c, creatinine, eGFR, triglycerides, total cholesterol, HDL cholesterol, lymphocyte count, liver function: ALT, ALP, albumin, bilirubin, BMI and blood pressure. Imputations were carried out separately for each medication development cohort. All predictors were included in the imputation modelling along with the cumulative hazard function and outcome indicator. 10 imputed datasets were created. Backwards elimination by means of the Akaike Information Criteria[10] to identify which of the predictors to retain in the final models was carried out for each imputed dataset for each medication cohort. Predictors were kept in the final model if they were retained in seven out of ten imputations[11, 12]. Final model parameters were obtained by combining estimates from the final model applied to each imputed dataset, using Rubin’s rules to adjust coefficients and standard errors to account for variability between imputations[13]. Levels of missingness are presented in Appendix table 1.

Stata (version 18) was used for these analyses.

***Model validation and comparisons***

To compute the validation statistics outlined in the Methods, missing data in the validation cohorts were imputed as for the development cohorts using multiple imputation, and each validation statistic computed for each of 10 imputed datasets and combined using Rubin’s rules[13]. In addition, an internal cross-validation within the development cohorts was carried out: the development cohorts were split into ten random “test sets”, each comprising equal numbers of individuals. A test set will be omitted from the cohort and the model re-built within the remaining nine test sets and then tested on the omitted test set. This was repeated excluding a different test set each time.

***Machine learning model development***

As outlined in the Methods, the following machine learning algorithms (all adapted for survival data) were considered: random forest[14], extreme gradient boosted model[15, 16], linear and kernel support vector machines[17], deep learning neural networks[18, 19] and a Super Learner ensemble, combining all the models together[20]. The Super Learner ensemble is a supervised learning method that uses a stacking process to determine the optimal weighted combination of a collection of the base machine learning algorithms[20]. Separate models were again developed for each medication development cohort, using the imputed data. The complete list of predictors were all included in the models. Python was used to develop the models. Python modules which adapt the machine learning algorithm to survival data were used, specifically: RandomSurvivalForest from sksurv.ensemble for the random forest; and GradientBoostingSurvivalAnalysis from sksurv.ensemble in scikit-survival for the extreme gradient boosted model[21]. FastSurvivalSVM from sksurv.svm in scikit-survival was used for the linear support vector machine, hyperparameter alpha (which determines the amount of regularization to apply) tuned via gridsearch (GridSearchCV), with 100-fold cross validations[21]. Likewise FastKernelSurvivalSVM from sksurv.svm in scikit-survival was used for the kernel support vector machine, with a radial basis function (RBF) kernel[21]. For the neural networks, we used CoxPH from pycox.models (also known as DeepSurv, from <https://nbviewer.org/github/havakv/pycox/blob/master/examples/01_introduction.ipynb>),[18, 19] in conjunction with PyTorch to define the neural net (MLPVanilla) and optimiser (Adam). Finally the different machine learning algorithms were combined as a SuperLearner ensemble following the methods in: <https://machinelearningmastery.com/super-learner-ensemble-in-python/>, adapted for survival data. The machine learning models performances in the validation cohort were compared with that of the Cox proportional hazards regression model, in terms of C statistics.

**Appendix table 1: Levels of missingness of incomplete predictor variables**

| **Development cohorts** | **Aripiprazole (n=904)** | **Olanzapine (n=2975)** | **Quetiapine (n=2408)** | **Risperidone (n=1968)** |
| --- | --- | --- | --- | --- |
|  | **Total non-missing (%)** | **Total non-missing (%)** | **Total non-missing (%)** | **Total non-missing (%)** |
| BMI, kg/m2 | 792 (87.6) | 2508 (84.3) | 2139 (88.8) | 1692 (86.0) |
| DBP, mmHg | 894 (98.9) | 2924 (98.3) | 2378 (98.8) | 1943 (98.7) |
| SBP, mmHg | 894 (98.9) | 2924 (98.3) | 2378 (98.8) | 1943 (98.7) |
| Total cholesterol, mmol/L | 776 (85.8) | 2486 (83.6) | 2047 (85.0) | 1707 (86.7) |
| HDL cholesterol, mmol/L | 745 (82.4) | 2413 (81.1) | 1970 (81.8) | 1661 (84.4) |
| Triglycerides, mmol/L | 685 (75.8) | 2230 (75.0) | 1812 (75.3) | 1525 (77.5) |
| HbA1c, mmol/mol | 647 (71.6) | 2041 (68.6) | 1643 (68.2) | 1367 (69.5) |
| Creatinine, µmol/L | 845 (93.5) | 2678 (90.0) | 2260 (93.9) | 1840 (93.5) |
| eGFR, ml/min/1.73m2 | 795 (87.9) | 2560 (86.1) | 2171 (90.2) | 1745 (88.7) |
| Lymphocyte count, 109/L | 848 (93.8) | 2675 (89.9) | 2251 (93.5) | 1837 (93.3) |
| Liver Bilirubin, µmol/L | 840 (92.9) | 2643 (88.8) | 2230 (92.6) | 1810 (92.0) |
| Liver Albumin, g/L | 841 (93.0) | 2643 (88.8) | 2232 (92.7) | 1818 (92.4) |
| Liver ALP, IU/L | 842 (93.1) | 2645 (88.9) | 2239 (93.0) | 1820 (92.5) |
| Liver ALT, IU/L | 721 (79.8) | 2257 (75.9) | 1878 (78.0) | 1500 (76.2) |
| **Validation cohorts** | **Aripiprazole (n=397)** | **Olanzapine (n=1306)** | **Quetiapine (n=1027)** | **Risperidone (n=826)** |
|  | **Total non-missing (%)** | **Total non-missing (%)** | **Total non-missing (%)** | **Total non-missing (%)** |
| BMI, kg/m2 | 341 (85.9) | 1097 (84.0) | 888 (86.5) | 700 (84.8) |
| DBP, mmHg | 391 (98.5) | 1281 (98.1) | 1022 (99.5) | 814 (98.6) |
| SBP, mmHg | 391 (98.5) | 1281 (98.1) | 1022 (99.5) | 814 (98.6) |
| Total cholesterol, mmol/L | 336 (84.6) | 1069 (81.9) | 862 (83.9) | 714 (86.4) |
| HDL cholesterol, mmol/L | 324 (81.6) | 1045 (80.0) | 828 (80.6) | 707 (85.6) |
| Triglycerides, mmol/L | 300 (75.6) | 977 (74.8) | 766 (74.6) | 661 (80.0) |
| HbA1c, mmol/mol | 288 (72.5) | 878 (67.2) | 695 (67.7) | 585 (70.8) |
| Creatinine, µmol/L | 365 (91.9) | 1180 (90.4) | 962 (93.7) | 768 (93.0) |
| eGFR, ml/min/1.73m2 | 347 (87.4) | 1132 (86.7) | 921 (89.7) | 740 (89.6) |
| Lymphocyte count, 109/L | 363 (91.4) | 1173 (89.8) | 970 (94.5) | 765 (92.6) |
| Liver Bilirubin, µmol/L | 358 (90.2) | 1162 (89.0) | 952 (92.7) | 760 (92.0) |
| Liver Albumin, g/L | 361 (90.9) | 1171 (89.7) | 952 (92.7) | 759 (91.9) |
| Liver ALP, IU/L | 362 (91.2) | 1167 (89.4) | 952 (92.7) | 759 (91.9) |
| Liver ALT, IU/L | 311 (78.3) | 1007 (77.1) | 785 (76.4) | 641 (77.6) |

**Appendix table 2: Distributions of predictor variables in the validation cohorts**

|  | **Aripiprazole (n=397)** | | **Olanzapine (n=1,306)** | | | **Quetiapine (n=1,027)** | | | **Risperidone (n=826)** | | |
| --- | --- | --- | --- | --- | --- | --- | --- | --- | --- | --- | --- |
|  | **Median** | **(IQR)** | | **Median** | **(IQR)** | | **Median** | **(IQR)** | | **Median** | **(IQR)** |
| Age at baseline, years | 34.9 | (25.3-51.8) | | 39.7 | (27.5-53.5) | | 38.2 | (27.1-53.3) | | 46.1 | (28.4-70.3) |
| Time from SMI diagnosis to baseline, days | 29.0 | (0-124) | | 20.0 | (0-67) | | 14.0 | (0-80) | | 21.5 | (0-85) |
| Number of hospitalisations in year before baseline | 0 | (0-1) | | 1 | (0-2) | | 0 | (0-1) | | 0 | (0-1) |
| Number of prior Elixhauser conditions | 2 | (1-3) | | 2 | (1-3) | | 2 | (2-3) | | 2 | (1-4) |
|  | **Mean** | **(sd)** | | **Mean** | **(sd)** | | **Mean** | **(sd)** | | **Mean** | **(sd)** |
| BMI, kg/m^2^ | 26.59 | (5.93) | | 24.8 | (5.11) | | 26.52 | (6.38) | | 26.34 | (5.98) |
| DBP, mmHg | 75.17 | (10.87) | | 75.74 | (10.67) | | 75.29 | (10.89) | | 75.14 | (10.9) |
| SBP, mmHg | 124.13 | (16.81) | | 124.71 | (16.59) | | 123.3 | (15.75) | | 127.09 | (16.6) |
| Total cholesterol, mmol/L | 4.88 | (1.16) | | 5.11 | (1.17) | | 5.01 | (1.16) | | 4.96 | (1.11) |
| HDL cholesterol, mmol/L | 1.39 | (.41) | | 1.41 | (.43) | | 1.4 | (.43) | | 1.43 | (.45) |
| Triglycerides, mmol/L | 1.57 | (1.15) | | 1.6 | (1.15) | | 1.61 | (1.2) | | 1.54 | (1.05) |
| HbA1c, mmol/mol | 41.85 | (23.59) | | 40.48 | (31.62) | | 38.9 | (18.4) | | 39.89 | (11.5) |
| Creatinine, µmol/L | 76.07 | (19.56) | | 77.45 | (17.98) | | 76.94 | (19.39) | | 80.97 | (38.72) |
| eGFR, ml/min/1.73m^2^ | 76.87 | (15.56) | | 78.52 | (15.43) | | 76.96 | (16.06) | | 75.22 | (18.12) |
| Lymphocyte count, 10^9^/L | 2.17 | (.74) | | 2.16 | (.94) | | 2.14 | (1.63) | | 2.12 | (1.49) |
| Liver Bilirubin, µmol/L | 8.81 | (5.61) | | 9.88 | (6.13) | | 9.26 | (5.6) | | 9.28 | (4.83) |
| Liver Albumin, g/L | 43.42 | (3.97) | | 43.2 | (4.37) | | 43.04 | (4.12) | | 42.6 | (4.47) |
| Liver ALP, IU/L | 84.59 | (44.2) | | 84.77 | (40.66) | | 80.99 | (38.01) | | 87.07 | (45.31) |
| Liver ALT, IU/L | 28.22 | (29.44) | | 29.09 | (27.86) | | 25.4 | (20.58) | | 25.86 | (21.27) |
|  | **N** | **(%)** | | **N** | **(%)** | | **N** | **(%)** | | **N** | **(%)** |
| Men | 163 | (41.1) | | 715 | (54.7) | | 381 | (37.1) | | 389 | (47.1) |
| **Ethnicity** |  |  | |  |  | |  |  | |  |  |
| Asian | 46 | (11.6) | | 99 | (7.6) | | 58 | (5.6) | | 64 | (7.7) |
| Black | 33 | (8.3) | | 96 | (7.4) | | 33 | (3.2) | | 54 | (6.5) |
| Mixed | 8 | (2) | | 24 | (1.8) | | 12 | (1.2) | | 28 | (3.4) |
| Other | 6 | (1.5) | | 30 | (2.3) | | 14 | (1.4) | | 16 | (1.9) |
| White | 304 | (76.6) | | 1057 | (80.9) | | 910 | (88.6) | | 664 | (80.4) |
| **IMD Deprivation quintile** |  |  | |  |  | |  |  | |  |  |
| 1 (least deprived) | 45 | (11.3) | | 190 | (14.5) | | 155 | (15.1) | | 118 | (14.3) |
| 2 | 65 | (16.4) | | 198 | (15.2) | | 169 | (16.5) | | 134 | (16.2) |
| 3 | 70 | (17.6) | | 236 | (18.1) | | 204 | (19.9) | | 132 | (16) |
| 4 | 95 | (23.9) | | 258 | (19.8) | | 216 | (21) | | 206 | (24.9) |
| 5 (most deprived) | 122 | (30.7) | | 424 | (32.5) | | 283 | (27.6) | | 236 | (28.6) |
| **SMI diagnosis** |  |  | |  |  | |  |  | |  |  |
| Bipolar | 104 | (26.2) | | 270 | (20.7) | | 543 | (52.9) | | 106 | (12.8) |
| Other psychosis | 245 | (61.7) | | 902 | (69.1) | | 443 | (43.1) | | 629 | (76.2) |
| Schizophrenia | 48 | (12.1) | | 134 | (10.3) | | 41 | (4) | | 91 | (11) |
|  | **N** | **(%)** | | **N** | **(%)** | | **N** | **(%)** | | **N** | **(%)** |
| Anxiolytics/ Hypnotics | 71 | (17.9) | | 276 | (21.1) | | 251 | (24.4) | | 139 | (16.8) |
| Antidepressants | 172 | (43.3) | | 566 | (43.3) | | 671 | (65.3) | | 329 | (39.8) |
| Prior depression | 208 | (52.4) | | 667 | (51.1) | | 703 | (68.5) | | 375 | (45.4) |
| Prior anxiety | 154 | (38.8) | | 459 | (35.1) | | 455 | (44.3) | | 288 | (34.9) |
| Prior personality disorder | 11 | (2.8) | | 32 | (2.5) | | 48 | (4.7) | | 26 | (3.1) |
| Prior eating disorder | 16 | (4) | | 24 | (1.8) | | 40 | (3.9) | | 13 | (1.6) |
| Prior PTSD | 5 | (1.3) | | 14 | (1.1) | | 26 | (2.5) | | 13 | (1.6) |
| Prior ACEs | 21 | (5.3) | | 63 | (4.8) | | 60 | (5.8) | | 42 | (5.1) |
| Prior Drug abuse | 29 | (7.3) | | 137 | (10.5) | | 90 | (8.8) | | 55 | (6.7) |
| Prior Alcohol abuse | 18 | (4.5) | | 118 | (9) | | 86 | (8.4) | | 60 | (7.3) |
| Atrial Fibrillation | 22 | (5.5) | | 37 | (2.8) | | 45 | (4.4) | | 60 | (7.3) |
| Diabetes | 138 | (34.8) | | 410 | (31.4) | | 353 | (34.4) | | 310 | (37.5) |
| Abnormal ECG | 18 | (4.5) | | 33 | (2.5) | | 47 | (4.6) | | 53 | (6.4) |
| Hypothyroidism | 18 | (4.5) | | 48 | (3.7) | | 48 | (4.7) | | 51 | (6.2) |
| Neurological disorders | 40 | (10.1) | | 96 | (7.4) | | 125 | (12.2) | | 116 | (14) |
| Liver disease | 3 | (.8) | | 33 | (2.5) | | 17 | (1.7) | | 20 | (2.4) |
| Renal disease | 24 | (6) | | 51 | (3.9) | | 49 | (4.8) | | 82 | (9.9) |
| HIV/ AIDS | 6 | (1.5) | | 21 | (1.6) | | 7 | (.70) | | 11 | (1.3) |

**Appendix table 3: Types of outcomes in each development and validation cohort**

Development cohorts

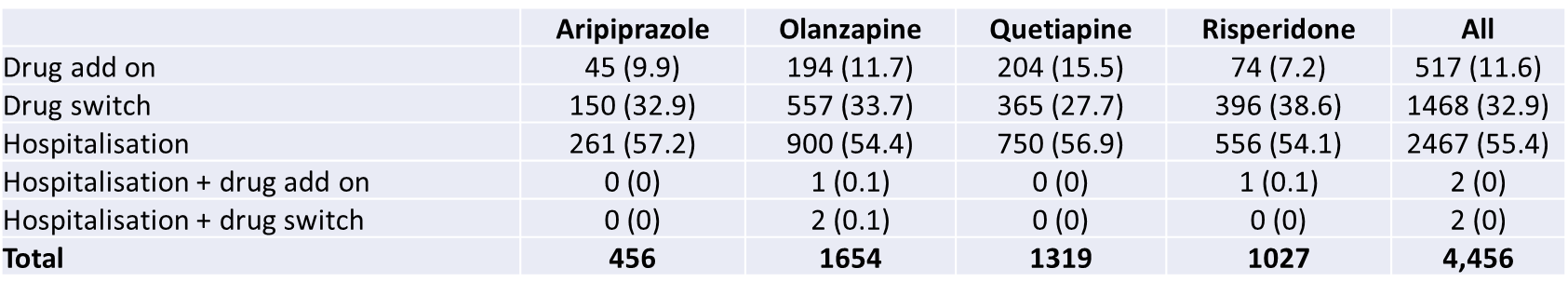

Validation cohorts

|  | **Aripiprazole** | **Olanzapine** | **Quetiapine** | **Risperidone** | **All** |
| --- | --- | --- | --- | --- | --- |
| Drug add on | 17 (7.9) | 73 (10.6) | 94 (16.2) | 48 (11.2) | 232 (12.1) |
| Drug switch | 63 (29.2) | 242 (35) | 164 (28.3) | 161 (37.4) | 630 (32.9) |
| Hospitalisation | 136 (63) | 375 (54.3) | 321 (55.4) | 220 (51.2) | 1052 (54.9) |
| Hospitalisation + drug switch | 0 (0) | 1 (0.1) | 0 (0) | 1 (0.2) | 2 (0.1) |
| **Total** | **216** | **691** | **579** | **430** | **1,916** |

NB no combined hospitalisation + drug add on in validation cohorts

**Appendix table 4: The five most common medication changes and add-ons in each development cohort**

Medication changes

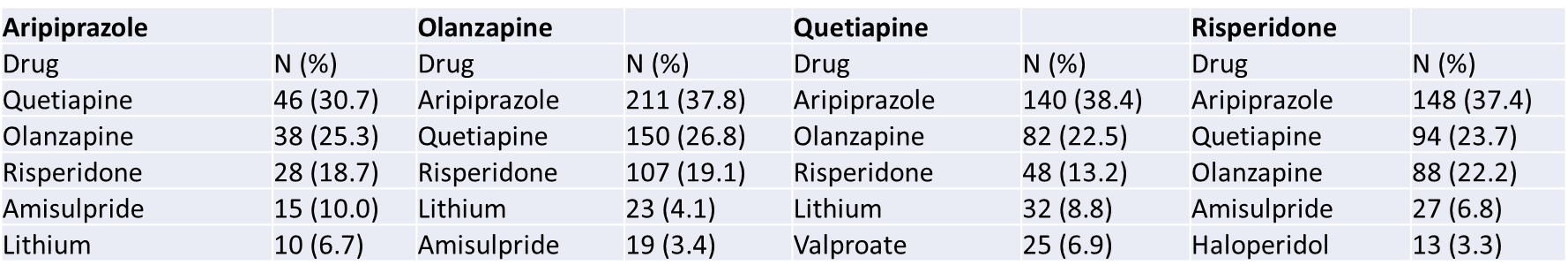

Medication add-ons

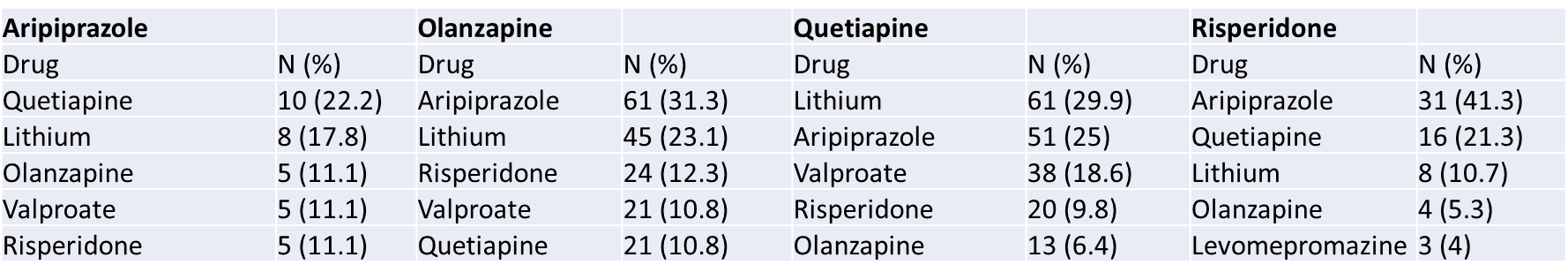

**Appendix table 5: Classical statistical prediction models after backwards elimination – starting from predictors totalling 23 coefficients**

Aripiprazole

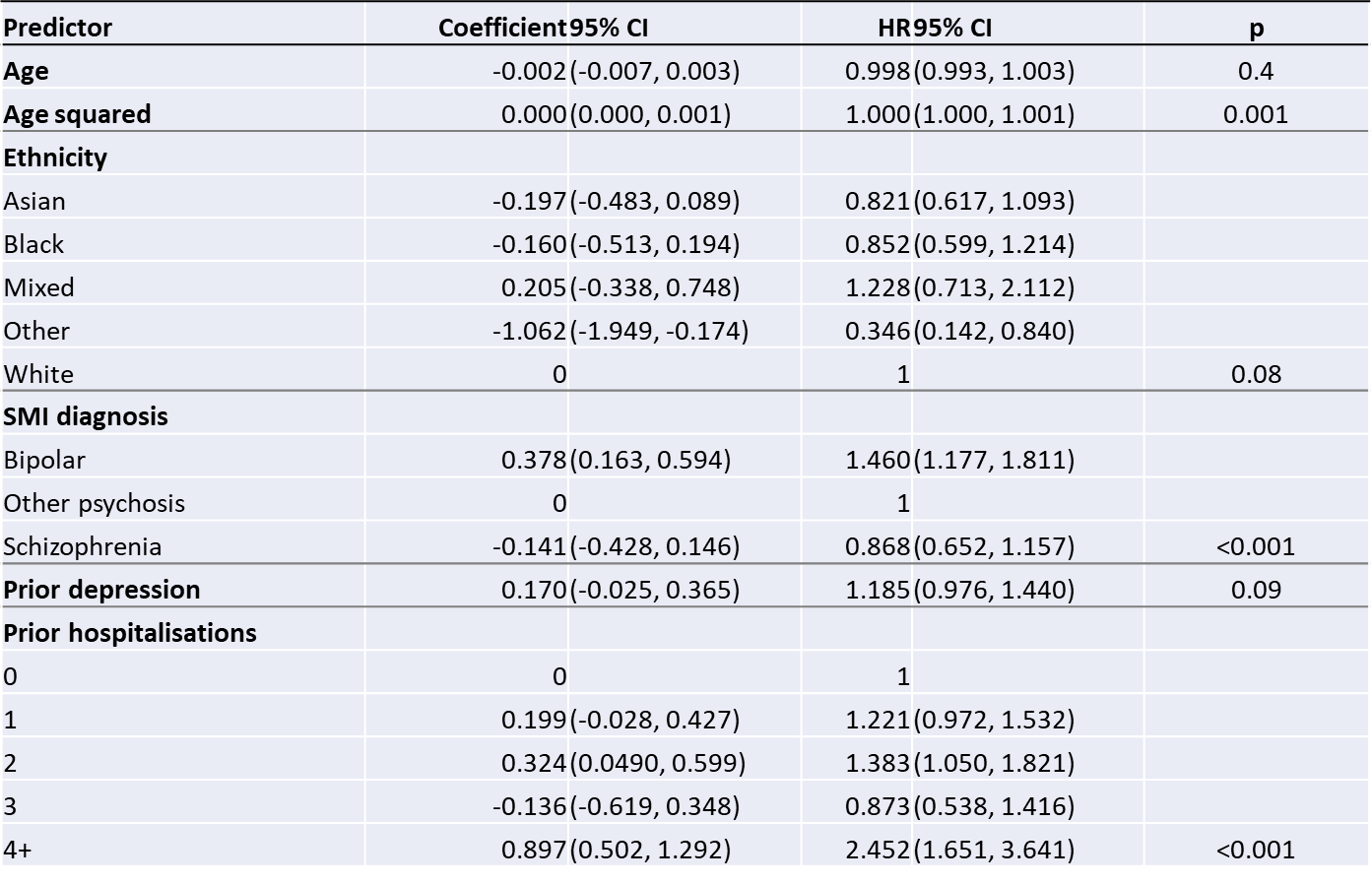

Olanzapine

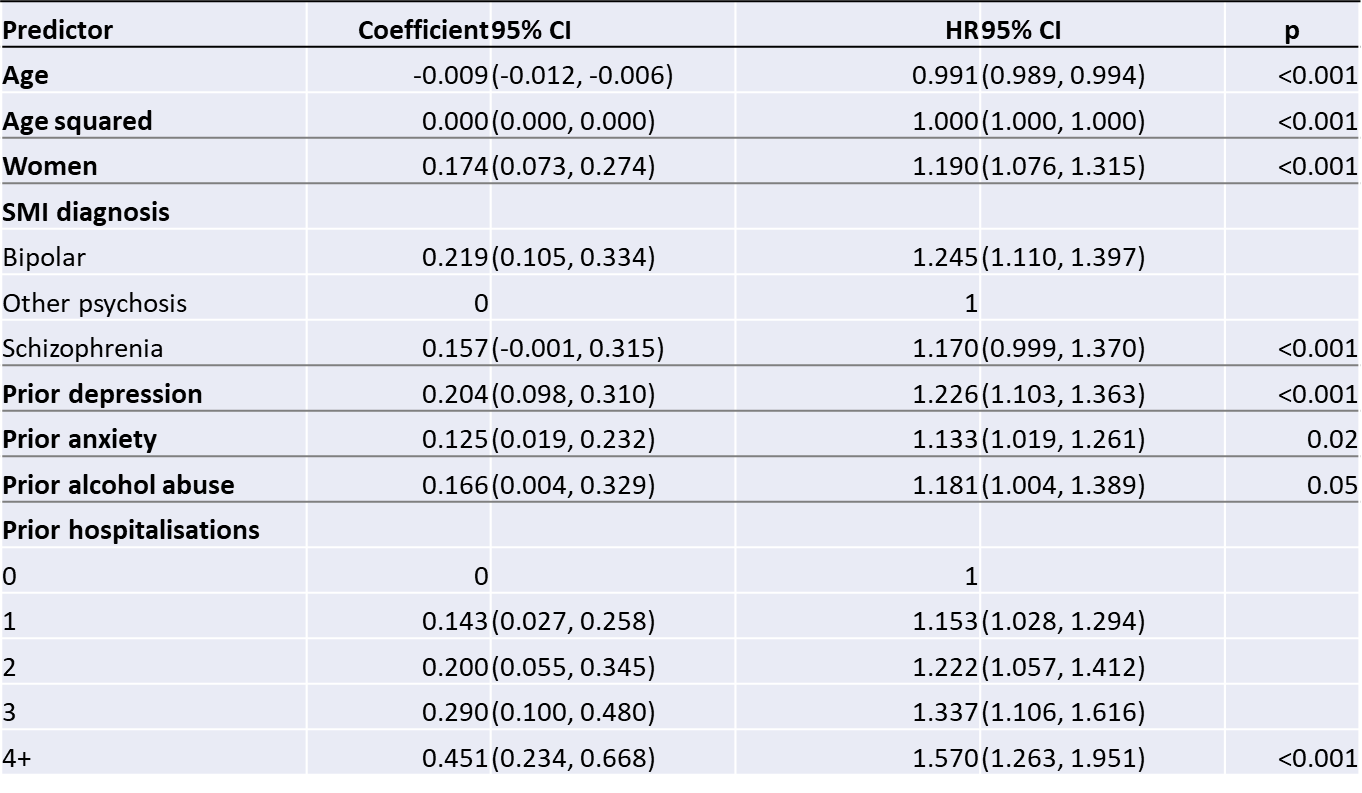

Quetiapine

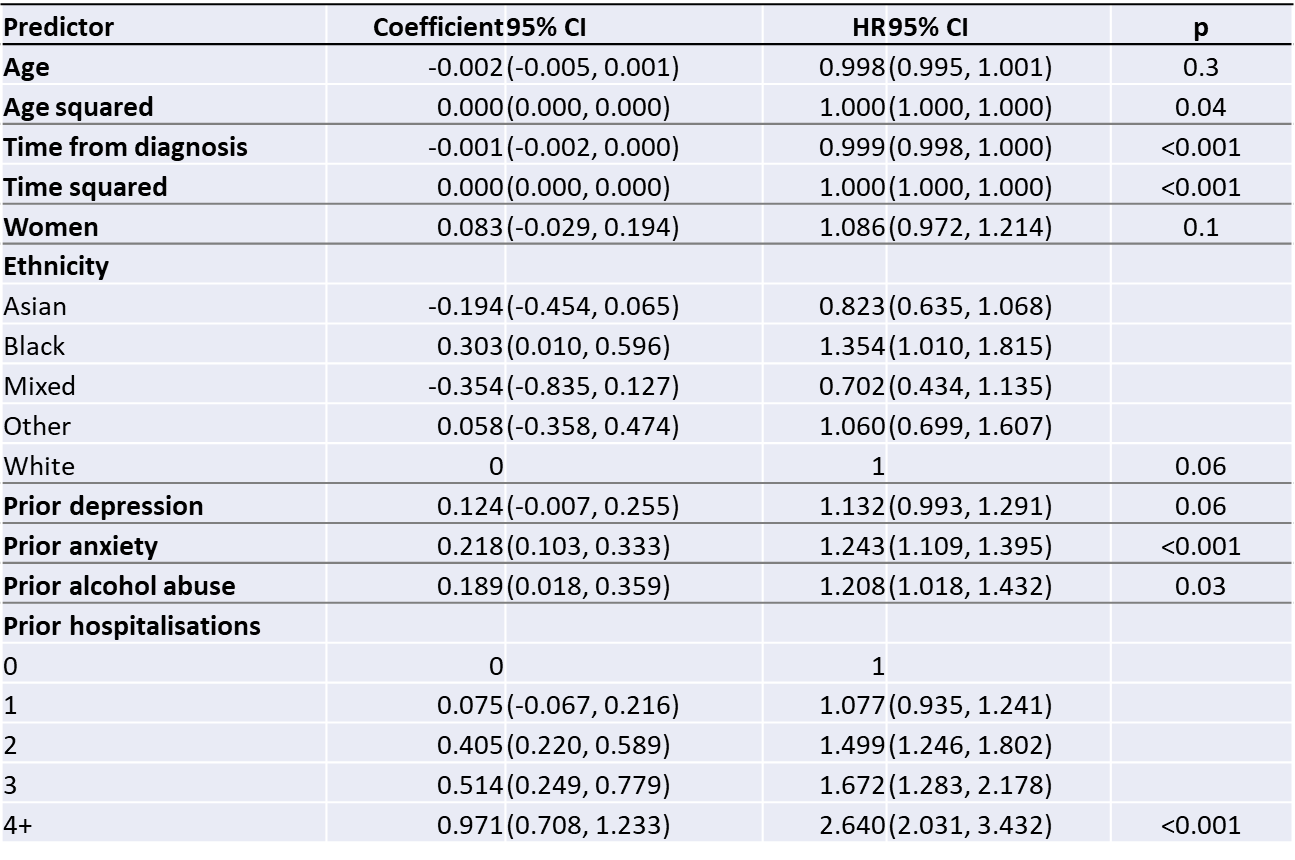

Risperidone

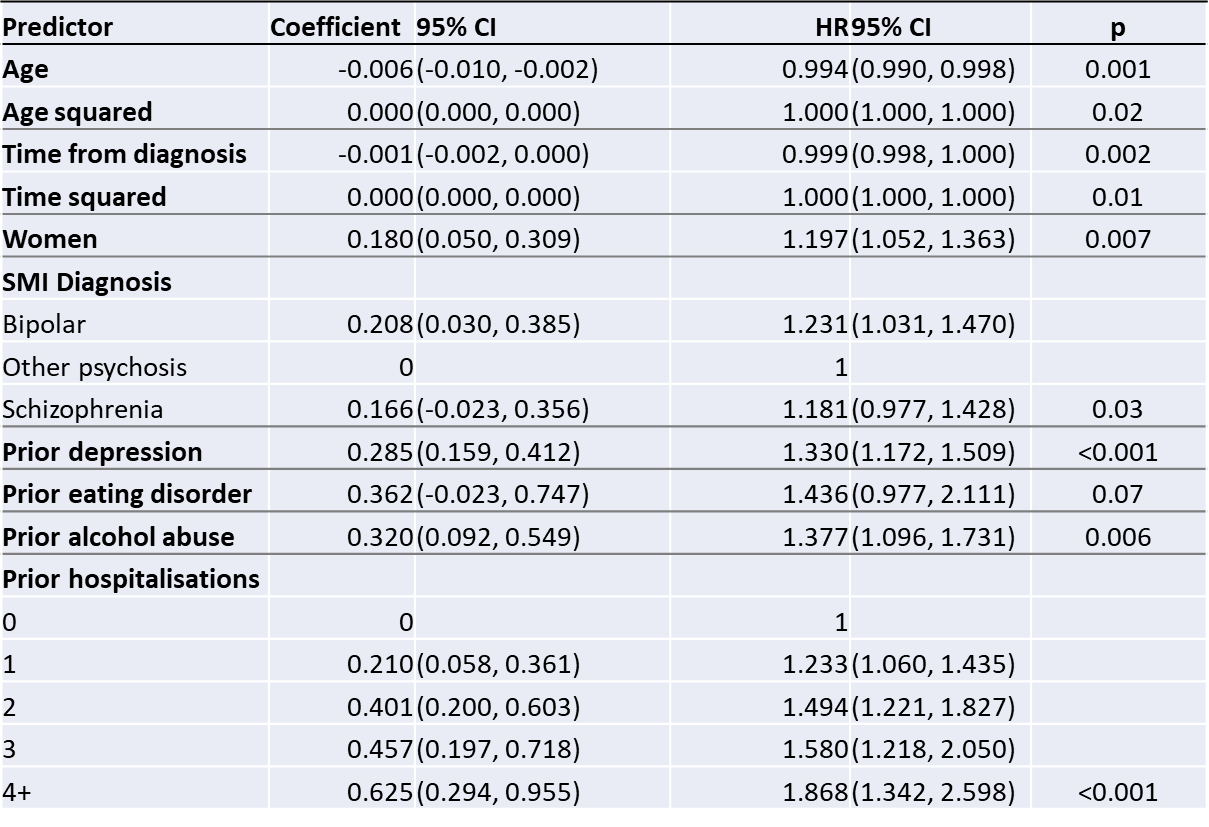

**Appendix table 6: Validation statistics for classical statistical risk prediction models**

|  |  |  |  | **Cut off: 55%** | | **Cut off: 60%** | | **Cut off: 65%** | |
| --- | --- | --- | --- | --- | --- | --- | --- | --- | --- |
| **Model** | **Discrimination: C (95% CI)** | **Discrimination: D (95% CI)** | **Calibration slope (95% CI)** | **Sensitivity** | **Specificity** | **Sensitivity** | **Specificity** | **Sensitivity** | **Specificity** |
| Aripiprazole key predictors | 0.56 (0.52, 0.60) | 0.31 (0.10, 0.52) | 0.49 (0.14, 0.84) | 52% | 61% | 40% | 73% | 21% | 85% |
| Olanzapine key predictors | 0.56 (0.54, 0.59) | 0.34 (0.22, 0.46) | 0.82 (0.54,1.10) | 90% | 14% | 76% | 36% | 55% | 55% |
| Quetiapine key predictors | 0.53 (0.50, 0.55) | 0.13 (0.002, 0.25) | 0.23 (-0.04, 0.49) | 85% | 21% | 65% | 41% | 44% | 60% |
| Risperidone key predictors | 0.61 (0.58, 0.63) | 0.67 (0.51, 0.83) | 1.28 (0.99, 1.57) | 87% | 27% | 71% | 48% | 53% | 64% |
| Aripiprazole all predictors | 0.57 (0.51, 0.63) | 0.30 (0.07, 0.53) | 0.22 (-0.04, 0.48) | 53% | 66% | 41% | 71% | 32% | 78% |
| Olanzapine all predictors | 0.54 (0.50, 0.57) | 0.35 (0.23, 0.47) | 0.67 (0.44, 0.90) | 74% | 35% | 58% | 51% | 40% | 69% |
| Quetiapine all predictors | 0.55 (0.52, 0.59) | 0.15 (0.02, 0.28) | 0.23 (0.01, 0.46) | 79% | 26% | 63% | 41% | 48% | 60% |
| Risperidone all predictors | 0.64 (0.59, 0.69) | 0.61 (0.45, 0.78) | 1.04 (0.74, 1.35) | 76% | 40% | 61% | 58% | 45% | 71% |

**Appendix table 7: Discrimination (C) statistics for machine learning risk prediction models compared with classical statistical risk prediction models**

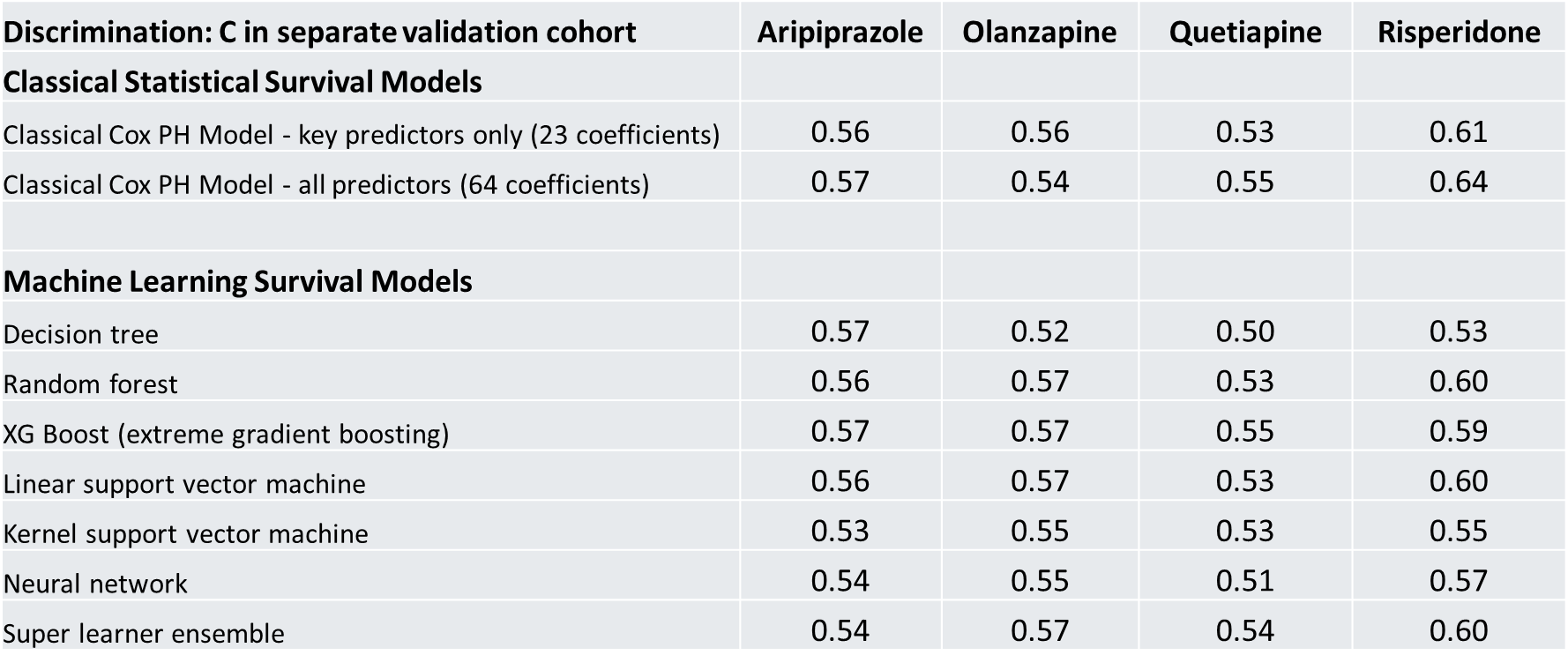
